# Fraud Prevalence and Prospective Prediction of Fraud Victimization in the Health and Retirement Study

**DOI:** 10.64898/2026.02.16.26346441

**Authors:** Melanie Leguizamon, Peter Lichtenberg, Laura Mosqueda, Emma Oyen, Belinda Y. Zhang, Daisy T. Noriega-Makarskyy, Cassidy P. Molinare, Jordan T. Williams, Jenna Axelrod, S. Duke Han

## Abstract

Financial exploitation of older adults is an increasingly prevalent public health concern, yet few have characterized fraud prevalence longitudinally or evaluated whether financial exploitation vulnerability measures prospectively predict fraud outcomes. Using data from the Health and Retirement Study, we examined fraud prevalence across a 14-year period and tested whether the Perceived Financial Vulnerability Scale (PFVS) predicts subsequent fraud victimization among older adults. Fraud prevalence increased steadily over time, rising from 5.0% in 2008 (347 of N=6,920) to a peak of 10.2% in 2022 (448 of N=4,380). Higher PFVS scores measured in 2018 were associated with greater odds of fraud victimization reported in 2022 (OR=1.62, 95% CI [1.25-2.15], *p*<.001). Most individuals who later reported fraud fell within the highest group of PFVS scores up to five years earlier. Together, these findings highlight financial exploitation as an emerging aging-related vulnerability and support the PFVS as a brief indicator of future fraud risk.

## Introduction

Financial exploitation of older adults is the most common form of elder abuse, increasing in prevalence as the population ages and resulting in profound personal and societal consequences^1^. Estimates suggest that, within a given year, between 5% and 17% of adults aged 60 years and older experience financial exploitation^2^, with national losses estimated at $28.3 billion annually^3^. More recently, financial exploitation has received increased attention within the field of psychology as an early risk factor for subsequent psychological^4^, social^5^, and cognitive decline^6^. As older adults may be uniquely vulnerable to financial exploitation due to age-related changes in wealth, cognition, decision-making abilities, and social connectedness^7^, it is critical to both characterize trends in fraud prevalence and identify reliable methods for determining who is most at risk for future exploitation.

Fraud victimization, a subtype of financial exploitation involving intentional deception for financial gain^8^, is frequently underreported by older adults and underestimated in the literature, in part due to varying exclusion criteria and measurement approaches^9^. As a result, fraud risk is often assessed using surrogate measures of scam susceptibility or perceived financial vulnerability. Commonly used instruments include the Susceptibility to Scams Scale^10^, the Assessment of Situation Judgment Questionnaire^11^, the Financial Exploitation Vulnerability Scale^12^, and the shortened, 6-item Perceived Financial Vulnerability Scale (PFVS)^13^. These measures have demonstrated good psychometric properties in community samples and are consistently associated with demographic, cognitive, and psychosocial correlates of vulnerability and well-being in older adulthood^14,15,16^.

Across studies, increased vulnerability to financial exploitation has been associated with older age, lower cognitive function, lower psychological well-being, and poorer health and financial literacy^17,18,19,20^. Beyond demographic and psychological factors, baseline wealth and changes in wealth over time have also been linked to perceived financial vulnerability^21^. However, because most of this work is cross-sectional, it remains unclear whether these correlates, and the vulnerability measures themselves, meaningfully predict future fraud risk.

The PFVS is particularly well-suited to clarify these issues given its brevity and ease of administration, applicability in longitudinal research, and demonstrated associations with factors implicated in fraud risk^21,22^. Importantly, perceived financial vulnerability may reflect broader agerelated changes in decision-making, cognition, and psychological well-being that precede future cognitive and functional impairment. Prior research has shown, for example, that indicators of psychological and functional vulnerability, including higher educational attainment and greater depressive symptoms, predicted subsequent fraud victimization over a four-year time frame^23^. Nevertheless, relatively few studies have examined fraud prevalence longitudinally, and none, to our knowledge, have established whether validated measures of perceived financial vulnerability prospectively predict fraud victimization in large population-based samples. The present study aims to address this gap by evaluating trends in fraud prevalence over time and testing whether PFVS scores prospectively predict future reported fraud victimization in a nationally representative longitudinal sample of older adults, thereby informing understanding of financial vulnerability in later life.

## Methods

### Participants

Participant data were obtained from the University of Michigan Health and Retirement Study (HRS), a longitudinal panel study funded by the National Institute on Aging (grant number U01AG009740) and the Social Security Administration. HRS data collection began in 1992, surveying adults aged 51-61 years and has followed participants every two years. Detailed information regarding the study design and data collection procedures has been published elsewhere^24,25^. As of the most recent 2022 data release, a total of 45,234 individuals across the United States had participated in at least one interview. Participants were first asked about fraud victimization in 2008. In 2018, a subsample of 1,400 participants were asked to complete the Perceived Financial Vulnerability Scale (PFVS). For the present study, fraud prevalence data were drawn from Waves 9 (2008) through 16 (2022) and PFVS data were drawn from Wave 14 (2018).

### Measures

#### Fraud Victimization

Beginning in 2008, participants completed an at-home Leave-Behind questionnaire^26^ following their core interview. This questionnaire assessed self-reported psychosocial issues and perceived limitations in the kind or amount of work a person could perform. Among these measures, two items assessed fraud victimization, defined here as a self-reported form of financial exploitation. Participants were asked whether they had been a victim of fraud in the past five years with response options of “yes” and “no.” Those who responded affirmatively were subsequently asked to report the year in which the fraud occurred. These questions were administered in waves 9, 10, 11, 15, and 16 (2008, 2010, 2012, 2020, and 2022, respectively) and were used as surrogate measures of fraud victimization in the current study. No additional questions were included regarding the type, frequency, or severity of the fraud.

#### Perceived Financial Vulnerability Scale

In the 2018 wave, 1,400 participants were asked to complete the PFVS. The PFVS was derived from 34 contextual items of the original 56-item Lichtenberg Financial Decision Rating Scale^27^. An initial 7-item PFVS was developed to assess financial awareness and psychological vulnerability related to personal finances^22^. Due to dichotomous wording and minimal variability, one item was excluded, resulting in a final 6-item measure. Items were rated on a 3-point Likert scale, yielding a total score ranging from 6 to 18, with higher scores indicating greater perceived financial vulnerability. The six PFVS items are presented in Table 3.

Normative validity testing of the PFVS demonstrated acceptable psychometric properties, including inter-item correlations ranging from 0.10 to 0.40, modest internal consistency (Cronbach’s α = 0.59), an adequate proportion of common variance (Kaiser-Meyer-Olkin value = 0.721), and a single factor best accounting for this variance (*Eigenvalue* = 2.0)^22^. In the present study, internal consistency was similarly modest but acceptable (Cronbach’s α = 0.56, 95% CI [0.52, 0.60])^28^.

#### Covariates

Covariate data were drawn from the RAND HRS Longitudinal file, which harmonized participant data across all survey waves from 1992 to 2022. Because covariates were drawn from the 2018 wave, use of this file ensured that covariate information was not missing due to variation in data collection across waves. Covariates included race, sex, years of education, and age and total household income measured at baseline (2018). Specifically, income was reported in nominal dollars and reflected the total household income for the previous calendar year. This measure was calculated as the sum of respondent and spouse earnings, pensions and annuities, Supplemental Security Income and Social Security Disability, Social Security retirement benefits, unemployment and workers’ compensation, other government transfers, household capital income, and other income sources.

#### Statistical analyses

Wave-specific fraud prevalence rates were calculated by identifying all participants who were asked and responded to the two fraud questions in each wave and dividing the number of individuals reporting fraud victimization in the past five years by the total number of participants with valid data for that wave. Logistic regression models were used to test whether fraud prevalence in later waves were significantly different from that observed in 2008.

Logistic regression analyses were also conducted to examine whether PFVS scores measured in 2018 prospectively predicted fraud victimization reported in 2022, adjusting for age, sex, race, education, and income. Due to small cell sizes in some race categories, race was coded as a binary variable using categories of White and non-White for regression analyses. Hispanic ethnicity was examined descriptively, but due to sparse cell counts and quasi-complete separation, it was not included as a covariate in regression models. Additionally, missing income values (n=58) were replaced with the sample median, and a binary indicator denoting missing income was included in the model. Because reports of fraud in 2022 could reflect victimization occurring prior to completion of the PFVS, participants who reported fraud occurring before 2018 were excluded. This restriction ensured temporal ordering such that PFVS scores preceded fraud outcomes and could be interpreted as prospectively predicting subsequent fraud victimization. Finally, PFVS scores were grouped into three categories to approximate sample-based tertiles and interpret fraud prevalence across low, medium, and high perceived financial vulnerability groups (scores of 6-7, 8-9, and 10-17, respectively). Sensitivity and specificity analyses, along with receiver operating characteristic (ROC) analyses, were conducted to evaluate the predictive validity of the PFVS for fraud outcomes.

## Results

The fraud prevalence sample was comprised of 31,324 participants across the five survey waves in which the fraud questions were administered. Specifically, 6,920 individuals responded in 2008, 8,167 in 2010, 7,252 in 2012, 4,605 in 2020, and 4,380 in 2022, with 34% of participants responding in more than one wave. Of note, the fraud items were not included in the 2016 or 2018 questionnaires. Among the 1,400 participants who were given the PFVS items in 2018, 315 were subsequently asked about fraud victimization in 2022. The final analytic sample consisted of 304 participants after excluding individuals who reported fraud occurring prior to 2018 and those with missing PFVS or covariate data. Respondents were, on average, aged 61.88 ± 9.23 years, and 58.22% were female. The sample was predominantly White/Caucasian (775.99%), with 11.84% identifying as Black/African American and 12.17% classified as Other. 13.82% of the sample self-identified as Hispanic. Participants had a mean education of 13.20 ± 2.92 years and a median household income of $51,522. On average, respondents scored 8.58 ± 1.90 on the PFVS in 2018, indicative of medium perceived financial exploitation vulnerability, and 6.25% reported fraud victimization in 2022 (Table 1).

**Table 1.** Sample characteristics.

| Variable | Final Sample (n = 304) |
| --- | --- |
| Age, years (mean ± SD) | 61.88 ± 9.23 |
| Sex, n (%) |  |
| Female | 177 (58.22%) |
| Male | 127 (41.78%) |
| Race, n (%) |  |
| White/Caucasian | 231 (75.99%) |
| Black/African American | 36 (11.84%) |
| Other | 37 (12.17%) |
| Ethnicity, n (%) |  |
| Hispanic | 42 (13.18%) |
| Non-Hispanic | 262 (86.18%) |
| Education, years (mean ± SD) | 13.20 ± 2.92 |
| Household Income, median | \$51,522 |
| Income missing, n (%) | 58 (19.08%) |
| PFVS Score (mean ± SD) | 8.58 ± 1.90 |
| Fraud Victimization, n (%) |  |
| Yes | 19 (6.25%) |
| No | 285 (93.75%) |

*Note.* Continuous variables are presented as mean ± standard deviation and categorical variables as counts and percentages. PFVS = Perceived Financial Vulnerability Scale.

Fraud prevalence increased across survey waves, with the highest prevalence (10.2%) observed in 2022 (Figure 1). Relative to 2008, the odds of reporting fraud victimization were higher in 2010 (OR = 1.20, *p* = .011) and 2012 (OR = 1.23, *p* = .005), and even greater in 2020 (OR = 1.69, *p* < .001) and 2022 (OR = 2.16, *p* < .001).

**Figure 1.**
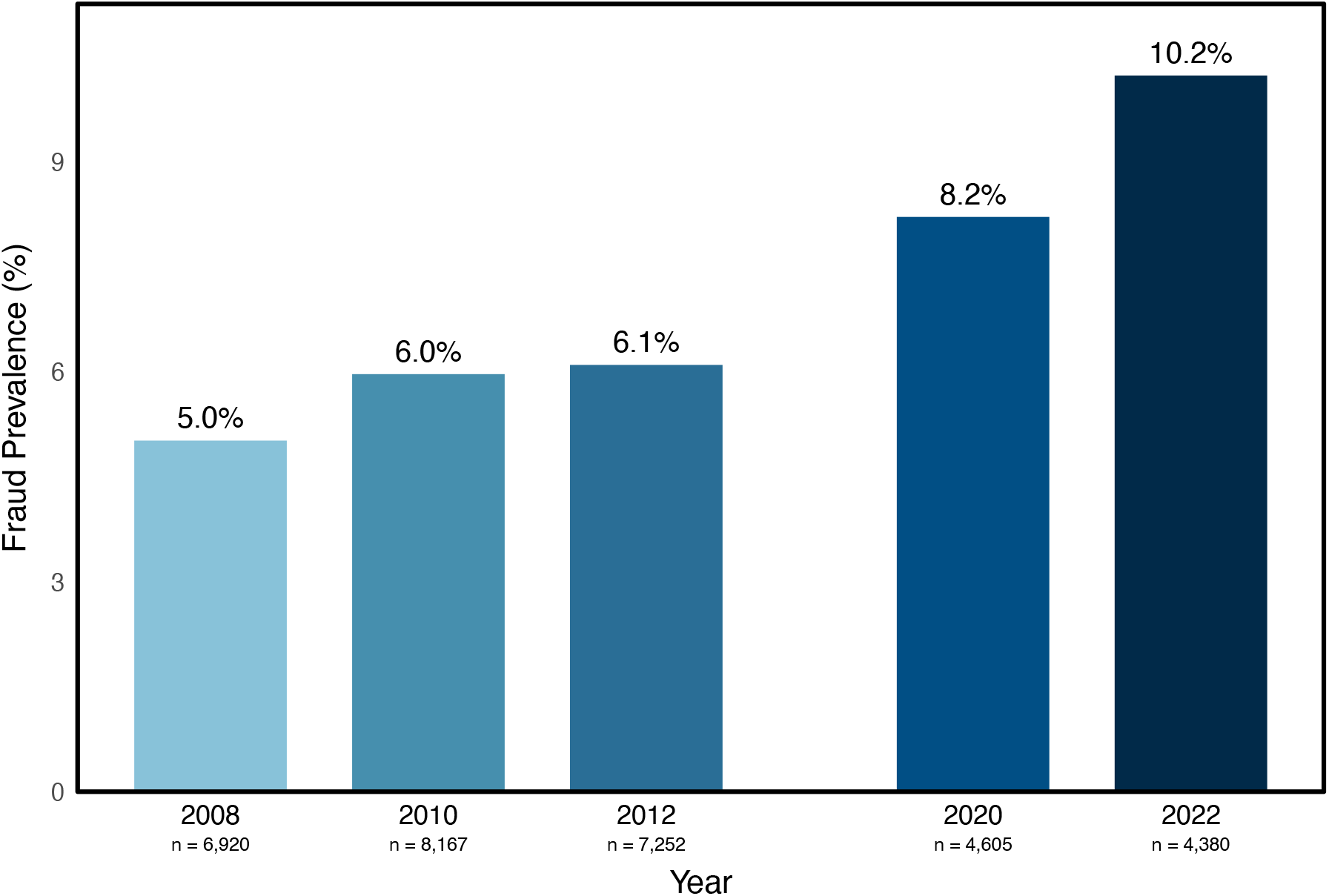
Fraud prevalence across waves.

Logistic regression analyses revealed that PFVS scores measured in 2018 were associated with increased odds of subsequent fraud victimization (OR = 1.62, 95% CI [1.25-2.15], *p* < .001; Table 2). Results remained unchanged in sensitivity analyses restricted to participants with complete income data (n = 246; OR = 1.63, 95% CI [1.22 – 2.26], *p* = .002). In contrast, demographic characteristics, including age, sex, race, education, and income were not associated with future fraud risk.

**Table 2.** Results of logistic regression models predicting fraud victimization reported in 2022 from Perceived Financial Vulnerability Scale score in 2018.

| Predictor | OR | 95% CI | p-value |
| --- | --- | --- | --- |
| Age (2018) | 1.01 | [0.96-1.07] | .654 |
| Sex (Female vs Male) | 1.38 | [0.51-4.03] | .535 |
| Race (Non-White vs White) | 1.80 | [0.41-1.62] | .701 |
| Education | 1.16 | [0.93-1.43] | .241 |
| Household Income (2018) | 1.00 | [1.00-1.00] | .779 |
| Income Missing | 0.65 | [0.13-2.51] | .561 |
| PFVS Score | 1.62 | [1.25-2.15] | **<.001\*** |

*Note.* Odds ratios (ORs) are extracted from logistic regression models predicting fraud victimization in 2022. Significant p-values are bolded with $*p < .001$ . PFVS = Perceived Financial Vulnerability Scale.

**Table 3.** Perceived Financial Vulnerability Scale survey items.

| Question Item | Question Text |
| --- | --- |
| 1 | How often do you feel anxious about your day-to-day financial decision transactions? Would you say never, sometimes, or often? |
| 2 | How often do you wish you had someone to talk to about financial decisions, transactions, or plans? Would you say never, sometimes, or often? |
| 3 | How worried are you that someone will take away your financial freedom? Would you say not at all worried, somewhat worried or very worried? |
| 4 | How confident are you in making big financial decisions? Would you say confident, unsure or not confident? |
| 5 | When it comes to making financial decisions and transactions, how often are you treated with less courtesy and respect than other people? Would you say never, sometimes or often? |
| 6 | How often has someone talked you into a decision to spend or donate money that you did not initially want to do? Would you say never, sometimes or often? |

For visual interpretation, PFVS scores were categorized into approximated sample-based tertiles (103 participants in the low group, 105 in the medium group, and 96 in the high group), corresponding to low (scores 6-7), medium (scores 8-9), and high (scores 10-17) perceived financial vulnerability. Among the full sample (n = 304), fraud prevalence increased across PFVS categories, rising from 1.9% in the low group to 4.8% in the medium group and 12.5% in the high group (Figure 2). Furthermore, of participants who experienced subsequent fraud, 63.2% scored in the highest PFVS category, followed by 26.3% in the middle category, and 10.5% in the low category (Figure 3A). Comparatively, among participants who did not experience fraud, PFVS scores were more evenly distributed, with 29.5% scoring in the highest category, 35.1% in the middle category, and 35.4% in the low category (Figure 3B).

**Figure 2.**
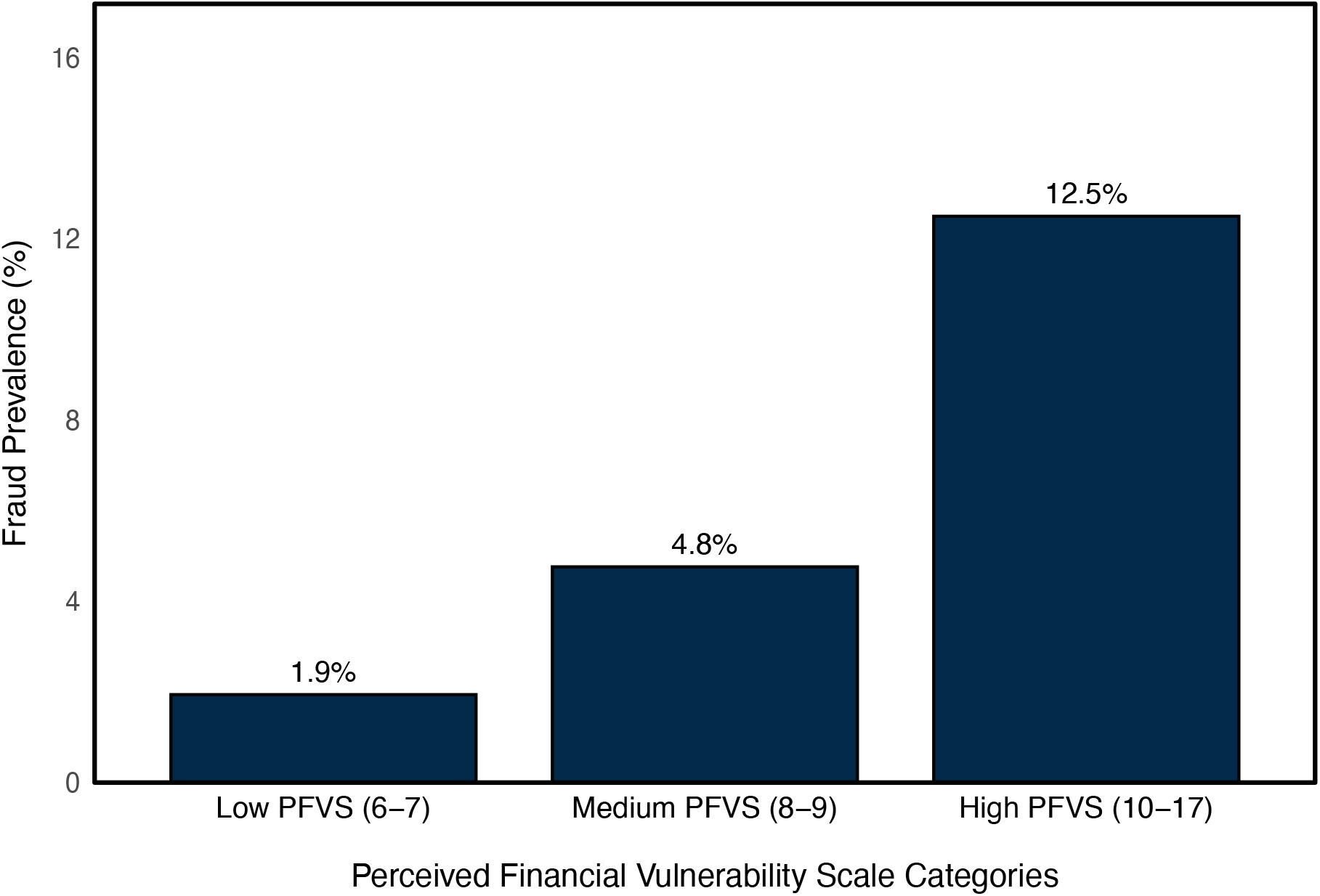
Fraud prevalence in 2022 across categories of Perceived Financial Vulnerability Scale scores measured in 2018. *Note*. Perceived Financial Vulnerability Scale scores were grouped into three categories approximating sample-based, with low = 6-7, medium = 8-9, and high = 10-17 points.

**Figure 3.**
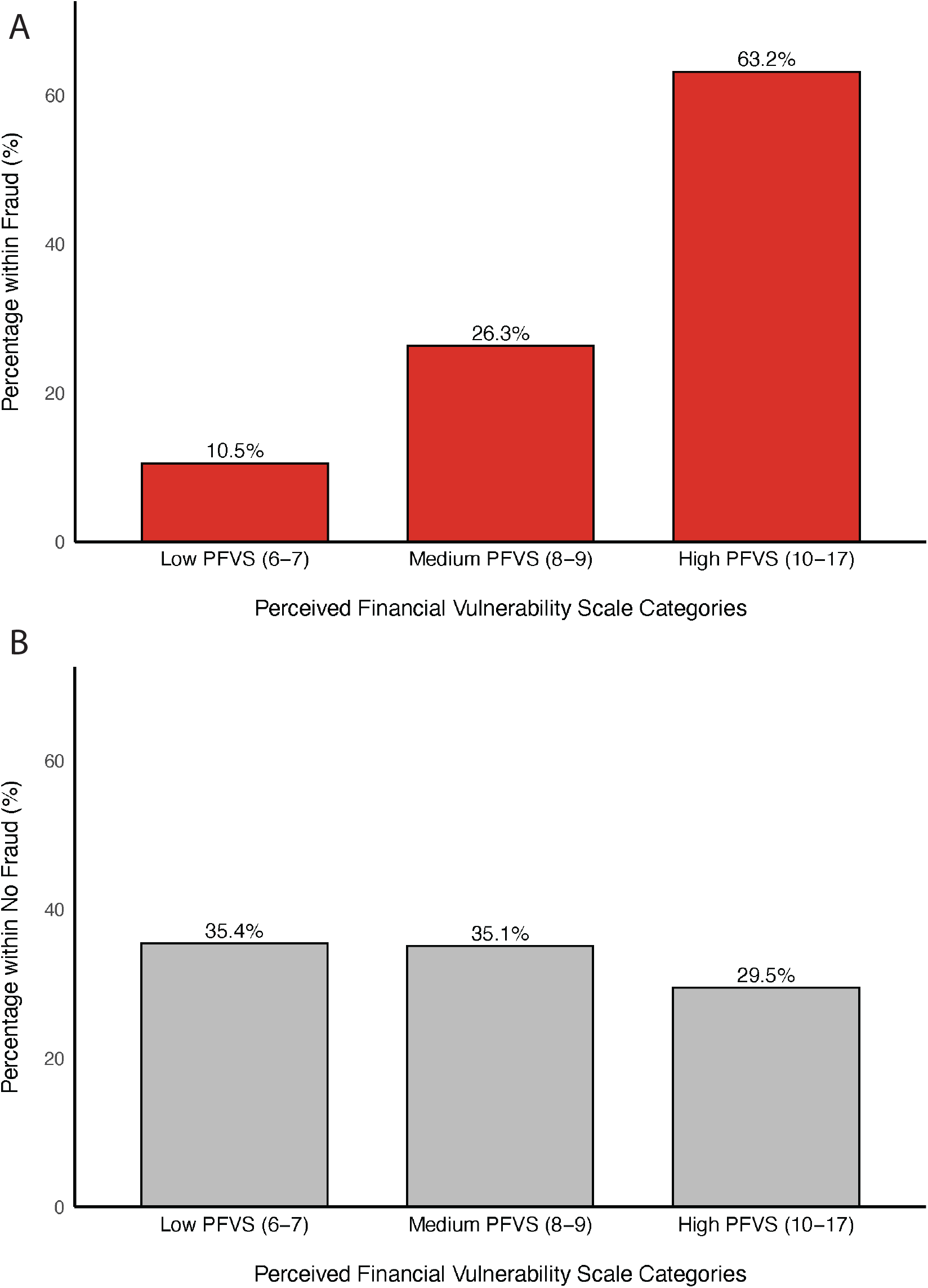
Distribution of Perceived Financial Vulnerability Scale categories among participants who experienced subsequent fraud victimization (A; n = 19) and those who did not (B; n = 285).

When PFVS scores were categorized into a binary higher-risk (highest category) versus lower-risk (lower two categories) group, the measure had moderate sensitivity (63%) and specificity (71%) for fraud victimization. Negative predictive value was high, indicating that individuals with lower PFVS scores had a 97% chance of not experiencing fraud. Positive predictive value was modest (13%), representing approximately a twofold greater risk of fraud among individuals with high PFVS scores, relative to the sample 6.25% base rate. Receiver operator characteristic analysis using continuous PFVS scores indicated acceptable discrimination (AUC = 0.70).

## Discussion

The present study provides empirical evidence in support of increasing fraud prevalence among older adults, highlighting financial exploitation as a significant real-world outcome associated with financial vulnerability in later life. In addition, our findings support the predictive validity of the PFVS as an indicator of future fraud risk. Across a 14-year period, fraud prevalence increased steadily, with the highest prevalence observed in 2022. Although fraud prevalence samples across waves were not fully independent (approximately 34% of participants contributed data to more than one wave), this pattern remains noteworthy given the observed doubling of fraud prevalence between the earliest and most recent assessments. Additionally, individuals who experienced fraud were more than twice as likely to score in the highest PFVS tertile compared with those who did not experience fraud. Overall, the PFVS demonstrated moderate sensitivity and specificity, as well as acceptable discrimination, for fraud victimization.

It has been widely suggested that fraud rates in older adulthood are likely to increase as the population ages and as new forms of fraud, including technology-based scams, become more prevalent^29^. Such changes are expected to lead to greater personal and societal consequences, which may impact financial security and individual autonomy. However, few studies have empirically documented changes in fraud prevalence over time using large, nationally-representative samples. Using data from the HRS, the present findings provide updated longitudinal evidence addressing this gap, underscoring the growing public health relevance of financial exploitation in older adulthood.

Previous research has linked the PFVS and related measures to factors associated with fraud risk, including age, psychological well-being, and financial literacy^18^, social connectedness and isolation^30,31^, cognitive function and impairment^32,33^, and neurobiological changes associated with neurodegenerative disease^34^. The present study advances this prior work by demonstrating that a validated measure of perceived financial vulnerability prospectively relates to subsequent fraud victimization. Although the value of subjective reports is often minimized relative to objective measures, which may be appropriate for some contexts, these findings highlight the importance of considering older adults’ perceptions of their own vulnerabilities when identifying individuals at heightened risk for fraud.

As susceptibility to financial exploitation has emerged as an early indicator of later-life impairment^6^, identifying such risk states at earlier stages may facilitate improved detection of individuals vulnerable to broader functional or cognitive decline and help prevent subsequent loss of financial resources. In our sample, participants with higher PFVS scores were at significantly greater risk of experiencing fraud in later years, with most individuals who reported fraud in 2022 having been classified in the highest PFVS group up to five years earlier. Extending prior cross-sectional work demonstrating associations between the PFVS and fraud-related vulnerability factors^21^, findings here provide longitudinal evidence that PFVS scores prospectively predict future fraud risk.

### Limitations

While this study contributes novel findings to the fraud and aging literature, several limitations should be acknowledged. Although the HRS was designed to be nationally representative and included a 2:1 oversampling of minority populations^35^, our sample nonetheless was comprised of a larger proportion of White/Caucasian participants relative to the U.S. population. Additionally, the use of three broad race categories (White/Caucasian, Black/African American, and Other) limited the ability to examine heterogeneity across specific racial and ethnic subgroups that may have unique vulnerabilities to fraud not fully captured by the PFVS. Furthermore, the wording of the fraud question included in the Leave-Behind questionnaire is vague and subject to individual interpretation. It is possible that respondents interpreted fraud victimization as a successful fraud attempt resulting in financial loss or as any attempt, including unsuccessful ones. Moreover, it remains unclear whether the approximately 2% increase in fraud risk observed between 2012 and 2020, representing a larger rise relative to prior wave comparisons, reflects the longer follow-up period. Alternatively, this rise may be attributed to increased exposure to fraud during the COVID-19 pandemic, possibly driven by social isolation and greater reliance on others^36^. Although a 2016 experimental module in the HRS assessed specific fraud types, including investment fraud, prize or lottery fraud, and account misuse, these items were not administered consistently across survey waves, precluding longitudinal analyses of fraud subtypes. Future research would benefit from more detailed and standardized measures of fraud experiences to differentiate between successful and unsuccessful fraud and to enable longitudinal assessment of specific fraud types.

### Surveillance Implications

Perceived financial vulnerability is not routinely assessed in clinical or community settings, despite its potential relevance for identifying individuals at heightened risk for future financial harm and possibly for detecting early vulnerability states that may precede later-life cognitive or functional decline. Findings from this study provide empirical evidence of increasing fraud prevalence among older adults in the United States and support the prospective validity of the PFVS as a brief indicator of fraud risk. Notably, even with only 19 reported fraud cases, elevated PFVS scores identified individuals at comparatively higher risk.

Thus, given its concise and efficient nature, the PFVS may be feasibly integrated into routine clinical or community assessments to evaluate fraud risk. It may also be applied in research and translational contexts to help identify older adults who could benefit from targeted education, increased monitoring, or preventative interventions aimed at reducing vulnerability to financial exploitation. Incorporating such measures might promote earlier detection of vulnerability and support efforts to preserve financial autonomy in later life. Further, clinicians and community leaders may play an important role in educating themselves, as well as their patients, caregivers, and community members, about the risks associated with financial exploitation vulnerability, particularly for those identified as being at elevated risk. More broadly, these findings underscore the importance of assessing financial exploitation vulnerability in later life and support the PFVS as a potential practical tool for characterizing risk in aging populations.

## Acknowledgements

This work was supported by the National Institute on Aging (K24AG081325 to SDH).

## Data Availability Statement

Data used in the present study is publicly available and can be found on the HRS website: https://hrs.isr.umich.edu/

